# High resolution proximity statistics as early warning for US universities reopening during COVID-19

**DOI:** 10.1101/2020.11.21.20236042

**Authors:** Zakaria Mehrab, Akhilandeshwari Goud Ranga, Debarati Sarkar, Srinivasan Venkatramanan, Youngyun Chung Baek, Samarth Swarup, Madhav V. Marathe

## Abstract

Reopening of colleges and universities for the Fall semester of 2020 across the United States has caused significant COVID-19 case spikes, requiring reactive responses such as temporary closures and switching to online learning. Until sufficient levels of immunity are reached through vaccination, Institutions of Higher Education will need to balance academic operations with COVID-19 spread risk within and outside the student community. In this work, we study the impact of proximity statistics obtained from high resolution mobility traces in predicting case rate surges in university counties. We focus on 50 land-grant university counties (LGUCs) across the country and show high correlation (PCC *>* 0.6) between proximity statistics and COVID-19 case rates for several LGUCs during the period around Fall 2020 reopenings. These observations provide a lead time of up to ∼3 weeks in preparing resources and planning containment efforts. We also show how features such as total population, population affiliated with university, median income and case rate intensity could explain some of the observed high correlation. We believe these easily explainable mobility metrics along with other disease surveillance indicators can help universities be better prepared for the Spring 2021 semester.

## 1 Introduction

Institutions of Higher Education (IHEs) are an integral part of a society’s normal operations. Like many other sectors, these have been adversely impacted by the COVID-19 pandemic, with many of the US universities moving academic operations entirely online [8], and/or requiring adaptations in pedagogy, thereby affecting student enrollment, access and engagement [14]. Many universities which opened up partly or completely in-person have subsequently seen case spikes leading to temporary closures. As of November 19, 2020, there have been up to 321,000 cases across 1700 colleges [16] spread across all 50 U.S. states. Most of these outbreaks subsequently led to case increases in the local community. In several cases, especially in rural counties, these contributed to a significant fraction of the overall cases to date. While comprehensive testing, tracing and isolation policies can and have brought some of these outbreaks under control, in many cases they have been reactive and ad hoc. Given the case surges seen across the country and worldwide as we head into the winter, questions of continued operations extending into Spring 2021 loom large in terms of COVID-19 control until the availability and mass roll-out of vaccines.

While standard surveillance measures such as pre-arrival testing [13], random prevalence sampling and innovative strategies such as wastewater surveillance can provide indicators of COVID-19 incidence in the student community, often these are lagging indicators of infection activity, hence have limited utility in resource management and outbreak containment. For effective response, identifying potential infection events will be valuable for public health officials in preparing for subsequent case rates and could also guide targeted surveillance and control measures. However, contact tracing based on mobile apps has been less effective so far, potentially due to low participation rates despite high acceptability [4]. As the next best alternative, one could use aggregate statistics derived passively from mobility traces obtained with consent to identify hotspots of interaction and potential subsequent case spikes. High resolution mobility data have become increasingly available and have been shown to be relevant for infectious disease modeling and forecasting across different phases [11, 6]. Anderson et al. [5] have also used such GPS traces to show the relationship to case spikes in some universities. While their focus was on understanding the impact of in-person instruction and inflow of students from across the country, the demographic characteristics of the region were not studied in detail. These will play a central role for local public health officials in understanding the potential impact on local community, and identify any leading indicators for triggering response efforts.

In our work, we use anonymized mobility traces to study their relationship to COVID-19 case rates at land-grant university counties (LGUCs) across the United States. Specifically, we focus on the level of proximity maintained by the users, as a proxy for social contact and disease spread risk. We show that simple, easily explainable proximity statistics computed from such mobility data serve as early indicators with high correlation (Pearson Correlation Coefficient, PCC *>* 0.6) with a lead time of up to ∼3 weeks, across multiple LGUCs. These results are especially evident in rural college towns and counties where the case spikes due to the student population are prominently observable at the county level. Further, demographic characteristics such as median income, university population partially explain the observed high correlation. Given the near real-time nature of such datasets (within 24 hours of interaction) and the privacy-preserving nature of aggregate statistics, they will serve as valuable early warning triggers for case investigation and resource planning by local health officials. In addition to providing a retrospective view of the Fall 2020 case spikes, they will be valuable for decision makers moving forward into the Spring 2021 semester.

## 2 Results

Our analyses show strong correlation between smoothed daily mean number of interactions and case rates in land-grant university counties (LGUCs) during the period from July to September 2020. Figure 1 (left panels) show the two curves for the top five LGUCs in terms of the largest maximum Pearson Correlation Coefficient (maxPCC) values observed across different lags (all 50 LGUCs are shown in Supplementary Figure S4). As evident from the visual comparison, strong spikes in interaction statistics are soon followed by case rate spikes in several of the LGUCs. The panels on the right side of Figure 1 show how the PCC varies with lead time along with the maximum PCC observed (vertical dashed line). They also show the results of Granger causality test at different lead times along with the level of significance achieved greater lead times (where the curve falls below the *p* = 0.05 level on the secondary y-axis).

**Figure 1:**
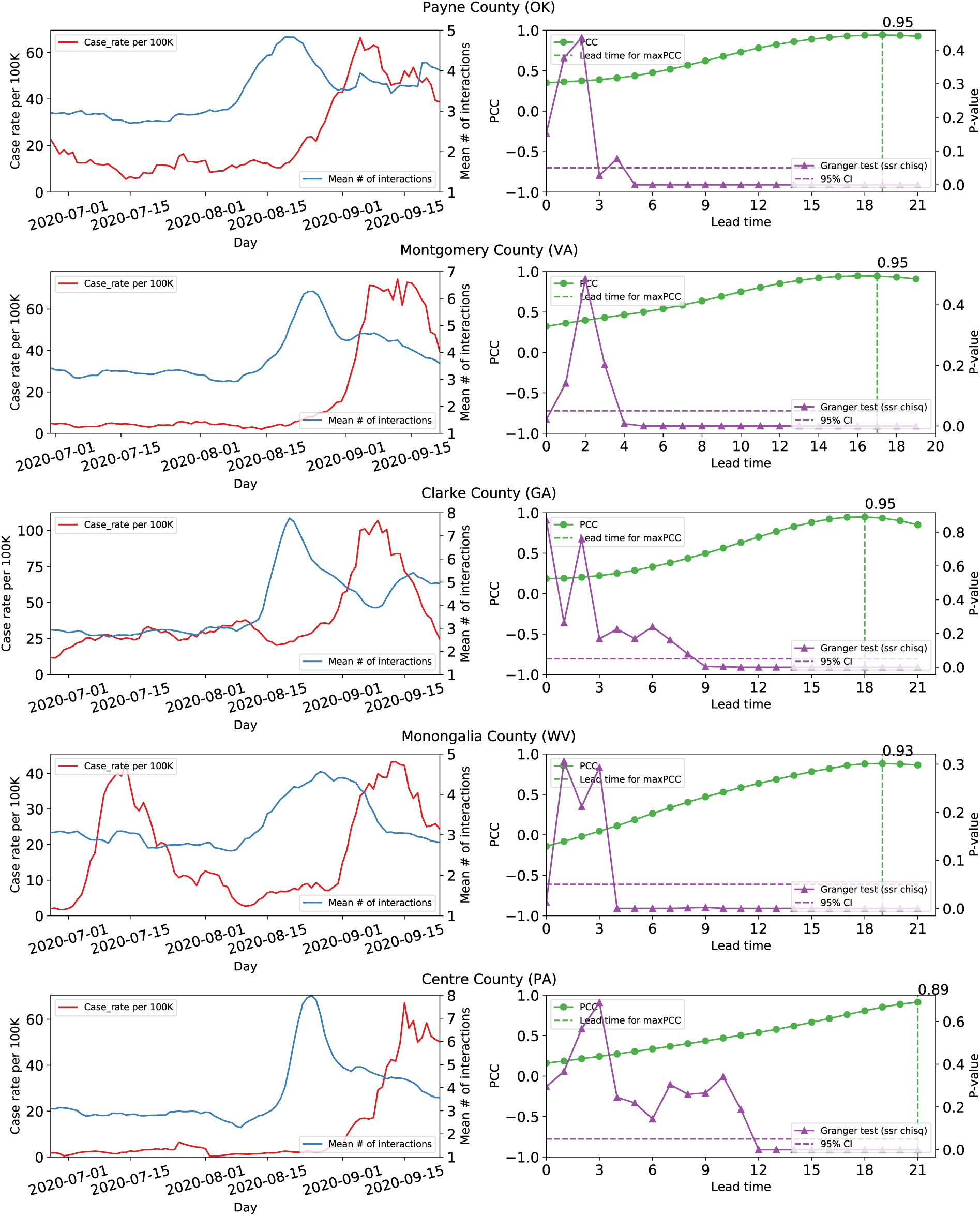
LGUCs with strong correlation. The name of the LGUC is given in the center-top position of each row. The left panel of each row shows the measured mean interaction (blue curve) and the measure case rate (red curve) for each day. We can see that a spike in the case rate is preceded by a spike in the mean interaction. For Monongalia County (WV), however, there is no preceding spike in interaction for the first peak in case rate. We believe this is due to boundary effect. The right panel show the PCC (green curve) and p-values (purple curve) for different lead times. PCC is high for lead times of 2-3 weeks and corresponding p-value is also significant across all the figures.

The maxPCC observed across different lead times is greater than 0.6 for more than half the counties under study, with the top three counties at maxPCC of 0.95. For all but three LGUCs, the maxPCC occurs at positive lead times strongly indicating causality (Figure 2). The median lead time for maxPCC is 14.5 days, and more than 80% of counties have maxPCC at a lead time of above 10 days. Further investigation using Granger causality reveals that for 68% of counties the measure is significant (*p <* 0.05, SSR chi-square test) for lead times above 10 days. The observed maxPCC values were also robust to hyperparameters of interaction statistics such as dwell time and minimum distance when computing proximity statistics (Supplementary Figure S2).

**Figure 2:**
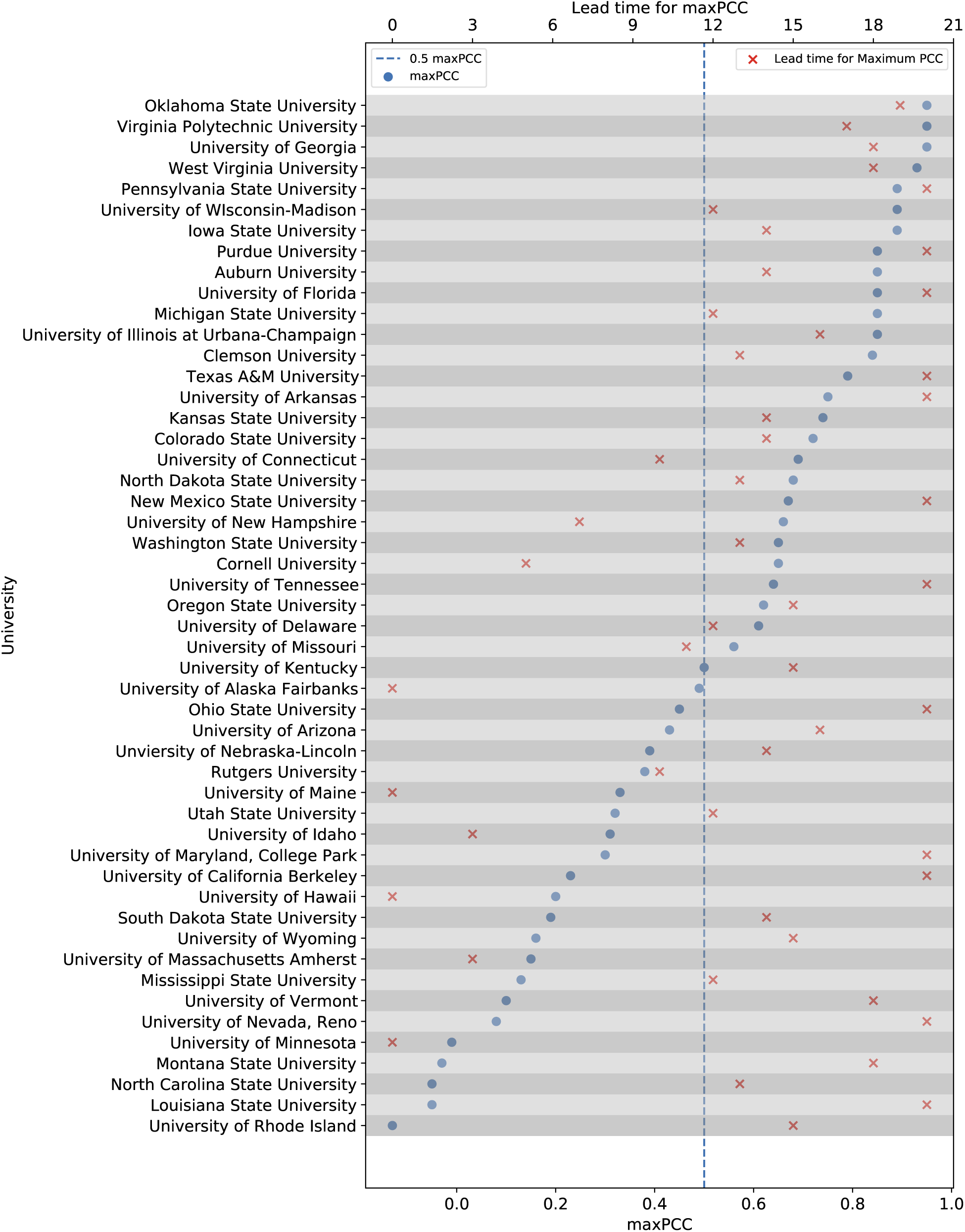
The maximum PCC and the corresponding lead time for each pair of interaction and case rate time series. We see that about half the university counties have maxPCC *>* 0.5 and lead time *>* 11.

Upon further investigation of the spatial and demographic characteristics, we notice that high correlations are mostly observed for LGUCs with smaller population sizes (blue circles in Figure 3). We also note that though these patterns are spread across the United States, especially in the Midwestern states and along the Eastern Seaboard, consistent with those regions experiencing higher COVID-19 incidence during July-September 2020. These patterns however are not universal, as seen in LGUCs with low maxPCC (Supplementary Table S2) or the corresponding lead time being negative (shown as 0 in Figure 2).

**Figure 3:**
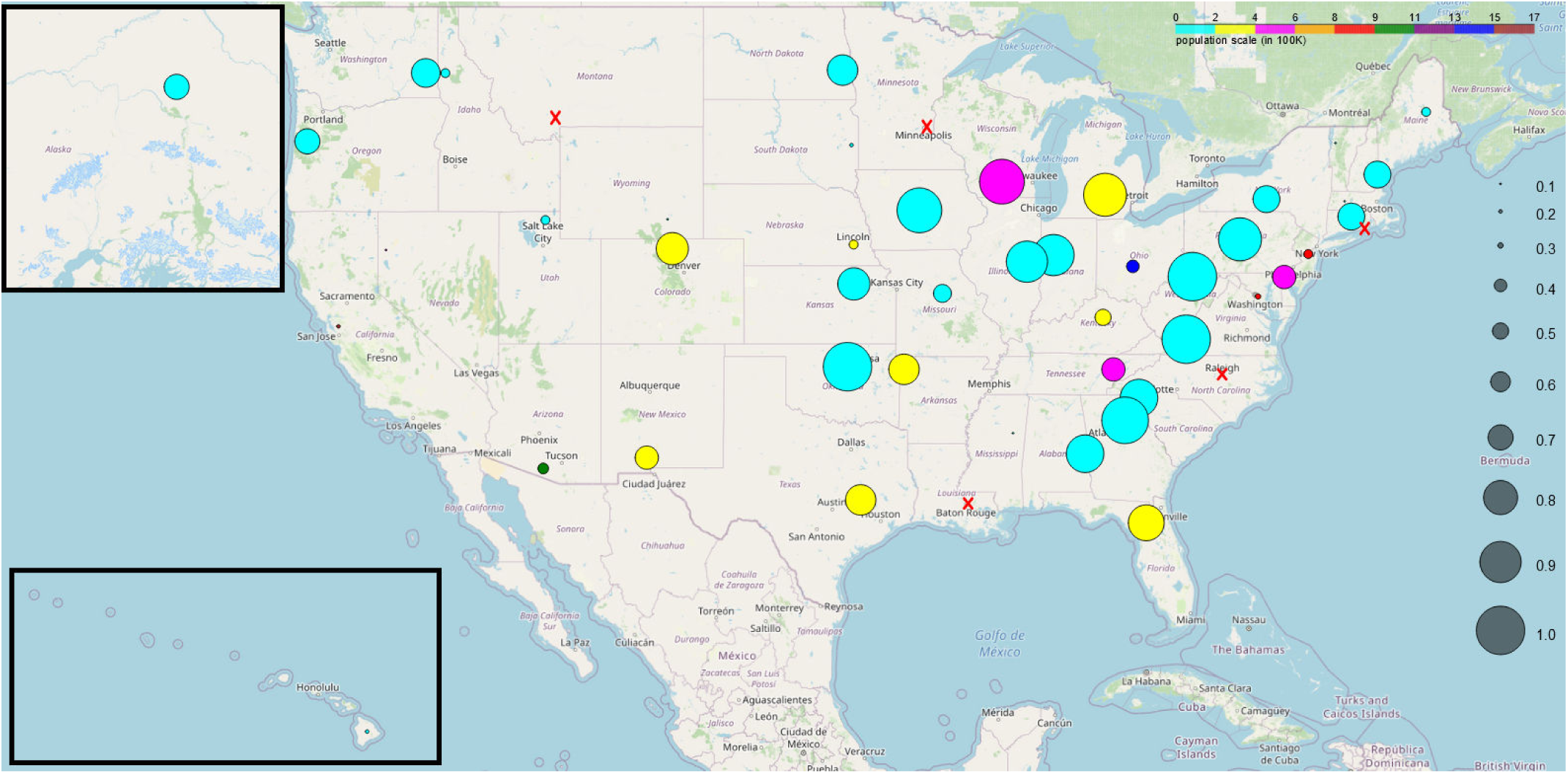
Population sizes and maxPCC values for LGUCs across the United States (insets contain Alaska and Hawaii). Circle size represents maxPCC. Larger circle represents high correlation in the corresponding LGUC. Also LGUCs with negative maxPCC are denoted with a cross (x) mark instead of circles. The color of the circle represents the population of the LGUC. We can see most of the large circles correspond to LGUC with low population (shown in cyan, yellow and magenta).

Regression models developed using various socioeconomic and epidemiological factors as independent variables to explain the maxPCC (either continuous or categorical) reveal that factors such as total LGUC population, number of people associated with university, rurality (captured by RUCC code of the county), median income level and epidemic intensity of case rate (measured as defined in [9]) were significant across multiple model variants (*R*^2^ ranging from 0.37 to 0.50). The chosen variables were mostly similar across different model selection procedures (forward, backward and stepwise regression) and dependent variable (continuous or categorical variant). A complete list of features considered and results for other model variants are provided in the Supplementary material.

## 3 Discussion

Human mobility datasets obtained from smartphones have been shown in the past to be useful for infectious disease modeling and forecasting. However, these suffer from demographic bias and may not be completely representative of the different population segments in the interaction counts, or in how they might be affecting case rates. In our analyses, we find that the correlation is highest in small college towns and weaker in larger, more urban, areas. Given the disproportionate representation of college-age individuals in the mobility data, their return to university campuses, especially rural ones leads to strong spikes in interactions. As shown in [5], their arrivals from higher incidence areas may have seeded outbreaks which subsequently caused case spikes observable at the county level. While a similar phenomenon could have played out in urban areas, given the background number of interactions these may be difficult to observe and thus fail to provide strong early warning signals for case spikes.

Based on the regression analyses, a positive coefficient associated with the university population depicts that maxPCC is higher in LGUCs where the interactions are attributable to a largely younger population. On the other hand, the overall LGUC population has a negative coefficient. This, coupled with the fact that university affiliated population is positively correlated with maxPCC, follows that maxPCC is high in small university towns; A negative coefficient associated with the Rural-Urban Continuum Codes (where higher values indicate lower urbanization) reflects the fact that many LGUCs are classified as metro areas. This is the case for all the counties in our data with the highest maxPCC values. The *median income* also has a negative coefficient, indicating that poorer counties show a higher maxPCC value. On a similar note, if there is a sufficient background case rate, then the change in case rate due to the change in interactions may not stand out sufficiently. In the regression analysis, the *entropy of the case rate* (computed as Shannon entropy of the normalized case time series, proposed by [9]) attempts to capture the strength of the signal in the case data, and is higher for counties that do not have noticeable surges in COVID-19 case rates. The coefficient for this feature is negative, which indicates that the stronger the spike the better the correlation with the interaction curve.

We note that other factors such as mask use prevalence, testing rates, and university operations (online vs in-person) may have an impact on whether high numbers of interactions lead to case spikes. However such datasets were not either universally available or did not match the timeframe of our study, so were not included. The goal of this study is to establish the utility of aggregate mobility statistics in creating early warning systems for universities. Thus it may not be exhaustive in explanatory variables and is not intended as a forecasting exercise. Further modeling of contact durations and disease transmissions using mechanistic models will definitely help improve the utility of such datasets. Operating on such anonymized datasets and computing aggregate yet useful statistics with privacy guarantees remains an active area of investigation. While most universities have tided over the case surges due to Fall 2020 reopening, the threat of subsequent outbreaks and impact on communities will persist until the rollout of a vaccine to the general population. Beyond retrospectively explaining case incidence in the past few months, this study can be seen as highlighting the value of proximity statistics as early warning indicators to case investigators and resource planners. As universities close early and plan their reopening strategies for Spring 2021, integrating such real-time indicators in their planning process will help with their response. We strongly believe that along with comprehensive testing and isolation plans, learning from the collective lessons of Fall experience, especially success stories, we could avoid similar spikes in the future despite surging case rates across the country.

## 4 Materials and Methods

### Datasets

We use data obtained via a third-party SDK operated by X-Mode [3] which serves multiple mobile applications and provides anonymized mobility traces with persistent identifiers (see Table S3 for statistics). We use TIGER/LINE shapefiles (for data extraction) [7] and medial income statistics [1] at county level from the US Census Bureau. County populations and COVID-19 case counts are obtained from USAFacts [2], while the RUCC codes were obtained from USDA [10]. Finally, we use a manually curated list of land-grant universities and their corresponding counties (Table S4). Additional details on all features considered are provided in the Supplementary material.

### Trajectory extraction

From the raw mobility traces obtained from X-Mode, we use Census shapefiles to first extract traces corresponding to the US and separate records per LGUC. We use a stop-point detection algorithm from scikit-mobility [12] to post-process the raw traces with a specified level of sensitivity (time interval 5 minutes, distance threshold 10m). Note that this step is prior to interaction computation, i.e., the time interval and distance threshold used in this step are distinct from the parameters used in interaction count computation below.

### Interaction statistics

We define an interaction as the co-occurrence of mobile device pings within a spatial distance threshold *d* and within temporal interval threshold *t*. Although more accurate interaction counts could hypothetically be obtained by interpolating travel to produce more continuous trajectories, the current method provides a lower bound on such interactions. Further, the use of stop points and subsequent deduplication allows us to obtain the number of unique interactions for each distinct user (i.e., device). We then compute the mean number of interactions per county. In our analysis, we set *d* = 2*m* and *t* = 1 *minute*. We also performed sensitivity analyses to these parameters (shown using Story County, Iowa as an example) and found that the results are robust to slight variations in *d* and *t* (Figure S2).

### Correlation analyses

We used two metrics, the Pearson Correlation Coefficient and Granger Causality, to measure the correlation between the obtained daily interaction statistics and case rates for the LGUCs of interest. Before analysis, county level case counts are converted to case rates per 100,000 and both case rates and mean interactions are smoothed by computing 7-day moving averages. The former is done to normalize across different population sizes, while the latter eliminates any within-week reporting artifacts and noise in trajectory or case data. PCC between the case rates and the mean interactions time series is obtained across different lead times and the maximum PCC and corresponding lead time are reported. We test Granger causality using python statsmodels [15] and report value for SSR-Chi square test.

### Regression analyses

We perform regression analyses for explaining the observed maxPCC values with various exogenous variables related to population characteristics and COVID-19 epidemiology. The features were normalized before the regression to account for the varying ranges of magnitude. We used three feature selection methods (forward, backward and stepwise) applied to continuous and quantized values of maxPCC to identify the statistically significant features. Among these models, we report the model obtained using the features obtained by backward selection. We find that the university population, total LGUC population, median household income, the RUCC type of the county and the entropy of case rate are strongly correlated with the maximum PCC (*R*^2^=0.504) (Table 1). A full list of the features, description of the feature selection methods and summary of all six linear regression models (Table S1) can be found in the Supplementary material.

**Table 1:** Linear regression analysis result for backward selection method and continuous target variable.

| Multiple $R^2$: 0.504; Adjusted $R^2$: 0.445 | | | | |
| --- | --- | --- | --- | --- |
| Variable | coef | std err | t | $P>\vert t\vert $ |
| Median Income | -0.9460 | 0.334 | -2.832 | 0.007 |
| University population | 0.8010 | 0.311 | 2.574 | 0.014 |
| Entropy of case rate | -0.6517 | 0.310 | -2.101 | 0.042 |
| Population | -0.0342 | 0.015 | -2.339 | 0.024 |
| RUCC Type | -1.2749 | 0.449 | -2.841 | 0.007 |
| const | 1.7834 | 0.448 | 3.979 | 0.000 |

## Data Availability

Aggregated statistics visualized in the manuscript are available upon request.

## Acknowledgement

The authors would like to thank members of the Biocomplexity COVID-19 Response Team and Network Systems Science and Advanced Computing (NSSAC) Division for their thoughtful comments and suggestions related to epidemic modeling and response support. We thank members of the Biocomplexity Institute and Initiative, University of Virginia for useful discussion and suggestions. This work was partially supported by National Institutes of Health (NIH) Grant R01GM109718, NSF BIG DATA Grant IIS-1633028, NSF DIBBS Grant OAC-1443054, NSF Grant No.: OAC-1916805, NSF Expeditions in Computing Grant CCF-1918656, CCF-1917819, NSF RAPID CNS-2028004, NSF RAPID OAC-2027541, US Centers for Disease Control and Prevention 75D30119C05935, DTRA subcontract/ARA S-D00189-15-TO-01-UVA. Any opinions, findings, and conclusions or recommendations expressed in this material are those of the author(s) and do not necessarily reflect the views of the funding agencies.

## Privacy and Ethics Statement

Anonymized datasets were obtained from X-Mode which collects location information from smartphone applications that use its software development kit (XDK). Users have the ability to limit/disable tracking and/or redistribution of their data. We refer the readers to https://xmode.io/xmode-privacy-policy-2/ for details on their Privacy Policy. The study at University of Virginia was approved by the Institutional Review Board for Social and Behavioral Sciences as part of the Human Research Protection Program. The paper only publishes aggregated derived statistics at the level of counties and on a daily basis guaranteeing individual privacy.

## Trajectory extraction and Stop point detection

**Figure S1:**
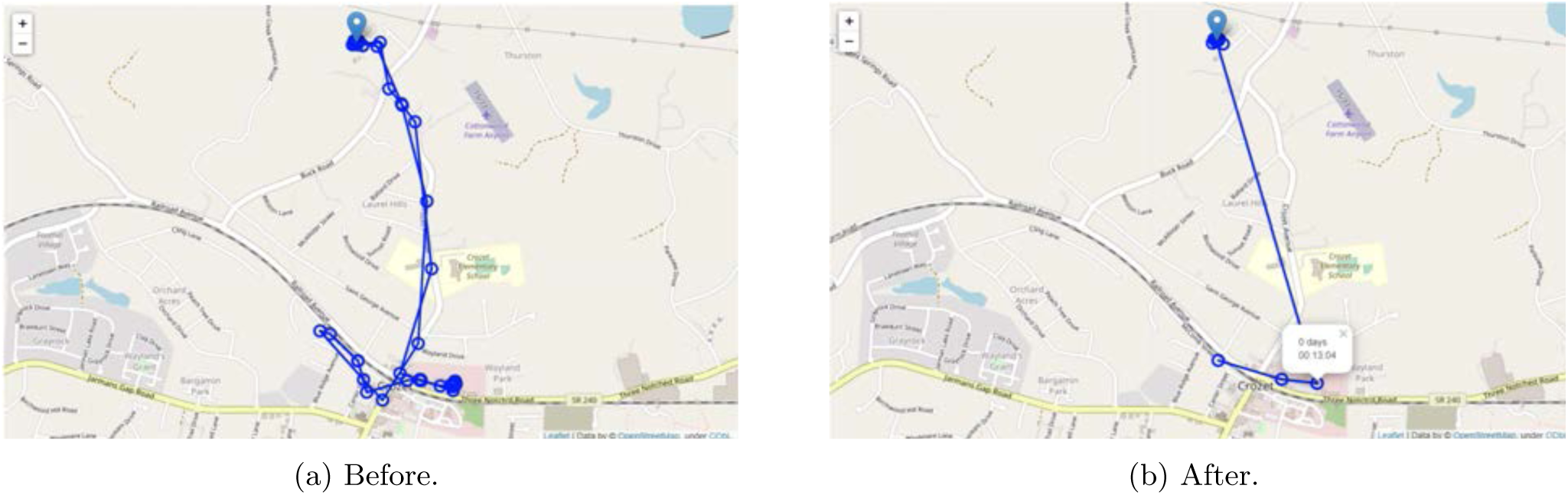
Effect of applying stop points.

For the purposes of our study the dataset spans 1 July, 2020 to 15 September, 2020 and focuses on mobility traces in the LGUCs of interest. From the mobility traces obtained for USA for a given day, we separate the pings by each LGUC. Then we use a stop point detection algorithm and generate a set of stop points from those raw trajectory points (Figure S1). The purpose of this pre-processing step is to filter out the records that do not contribute to significant interaction. For instance, this eliminates the spurious pings generated in transit and provide a ‘cleaner’ trajectory with fewer records for subsequent calculations. This also helps remove users who did not spend significant time within the county, but were transiting through the region.

We use the stop-point detection method implemented in the scikit-mobility [5] library. The stop point detection method requires one temporal constraint and two spatial constraints. The temporal constraint specifies the minimum number of minutes an individual has to spend at a location for that record to be considered a stop point. The two spatial constraints are stop radius factor and spatial radius. The product of these two parameters gives us a distance threshold. Any point from the candidate trajectory point within this distance threshold will be considered as a single stop point and the final stop point coordinates are the median latitude and longitude values of the points found within the specified distance threshold. We use 5 minutes as our temporal constraint. For the spatial constraints, we set spatial radius to 0.2 km and the stop radius factor as 0.05. Thus the distance threshold is 10 meters.

**Figure S2:**
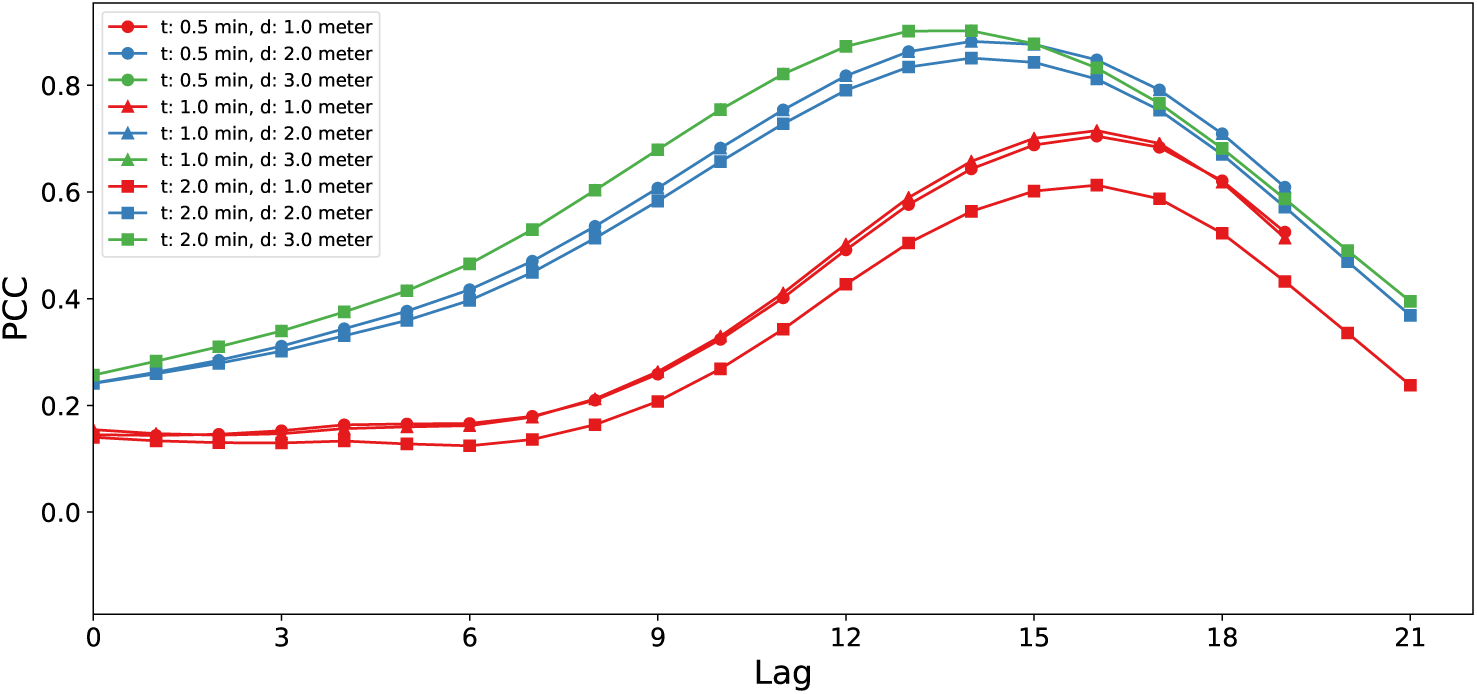
Sensitivity of Iowa State University LGUC PCC analysis with respect to the distance and time interval parameters.

## Measuring Interaction rate

After performing pre-processing on the previous step, we estimate the number of interactions occurring on each day for each county. Human interaction is associated with both temporal and spatial proximity. Intuitively, an interaction occurs when two persons are present close to each other (spatial) simultaneously (temporal). Following this, we define that an interaction happens when two people are present at a location within a spatial threshold *d*, and the difference of their time of pings is within a temporal threshold *t*. The data for a county for each day consists of a set of pings *P*. The set of pings are the stop points obtained from the previous step. Each ping *p*_*i*_ of *P* is defined by a tuple as follows: *p*_*i*_ = (*u*_*i*_, *lat*_*i*_, *lon*_*i*_, *ts*_*i*_). Here, *u*_*i*_ denotes an anonymous user. Although one user may have multiple *u*_*i*_ if he/she possesses multiple devices, for our experiment we consider each distinct *u*_*i*_ to be a separate user. *lat*_*i*_ and *lon*_*i*_ represent the latitude and longitude of the ping, respectively, and *ts*_*i*_ denotes the timestamp when the ping was captured. From the pings collected for each day, we calculate the number of unique interactions for each distinct *u*_*i*_. After calculating the number of unique interactions for each distinct user *u*_*i*_, we obtain the mean interaction count for that day for the particular county.

## Correlation Measurement

After obtaining the time-series data of interaction for a county, we aim at measuring whether any correlation exists between these data and the case rate data. As mentioned in the main text, correlation analyses are performed on 7-day smoothed versions of the mean number of interactions per county and the case rates per 100,000.

## Pearson Correlation

This is a statistic that measures the correlation between an exogenous variable *X* and an endogenous variable *Y*. It takes *X* and *Y* as input and generates a value between +1 and -1. A value close to 1 represents a strong positive correlation between *X* and *Y* (*Y* increases as *X* increases). A value close to -1 represents a strong negative correlation (*Y* decreases as *X* increases). A value of 0 indicates that there is no correlation between *X* and *Y* .

For our purposes, *X* is the time series of interactions and *Y* is the time series of case rates. We shift *X* by some lead time and compute the Pearson correlation coefficient for each such lead time. This not only tells us whether *X* and *Y* are correlated, but also gives us the optimal lead time that lets us predict *Y* from *X* if they are strongly correlated.

## Granger Causality Analysis

The Granger causality test is a statistical hypothesis test for determining whether one time series is more useful in forecasting another time series than the past values of the other time series itself. A time series *X* is said to “granger-cause” *Y* if it can be shown that the past values of *X* along with past values of *Y* provide more statistically significant prediction of the future value of *Y* than the future value of *Y* obtained from past values of *Y* alone. This is done through a series of chi square-tests and F-tests.

## Linear regression

The following features were considered in linear regression for predicting maxPCC of each county. For each feature, we first define what the feature represents and also explain how the feature was pre-processed for regression analysis, wherever applicable.

1. Standard deviation scale of interactions: The ratio of the standard deviation to the mean for the interaction rate time-series data. (Feature was normalized before usage)
2. Standard deviation scale of cases: The ratio of the standard deviation to the mean for the case rate time-series data. (Feature was normalized before usage)
3. Maximum standard deviation of interactions. (Feature was normalized before usage)
4. Maximum standard deviation of cases: (Feature was normalized before usage)
5. Income: Median household income for the year 2018 of the county as per the data provided by the Economic Research Service, USDA [1]. (Median Income values were in 100K range. So, the values were used on a scale of 100K in the regression analysis.)
6. Dynamic range of case rates: The difference between the maximum and minimum value in the case rate time series data. divided by the minimum value of the case rate time series data. (Feature was normalized before usage)
7. Dynamic range of interaction rates: The difference between the maximum and minimum value in the interaction rate time series data divided by the minimum value of the interaction rate time series data. (Feature was normalized before usage)
8. Entropy of case rates: The amount of uncertainty in the time series data for the case rate. This was computed by quantizing the values of the case rate time series data, converting it into a probability distribution and computing the entropy of the probability distribution.
9. Entropy of interaction rates: The amount of uncertainty in the time series data for the interaction rate. This was computed by quantizing the values of the interaction rate time series data, converting it into a probability distribution and computing the entropy of the probability distribution.
10. Expected age group: Weighted average of the age group of people in the county as per the data of 2019 provided by the US Census Bureau [2]. (Feature was normalized before usage)
11. University population: The number of people affiliated with the corresponding land grant university of the county as per IPEDS [4]. (On a scale of 100K)
12. Population: Population of the county (On a scale of 100K)
13. University percentage: Ratio of 11 to 12.
14. User coverage: The number of unique users in our dataset. (On a scale of 100K)
15. User coverage percent: Ratio of 14 to 12.
16. Social distancing index: Average social distancing index of a county as provided by University of Maryland [6]. (Feature was normalized before usage)
17. Type: RUCC Type of a county, stating whether a county is urban or rural. This is obtained from United States Department of Agriculture [3]. (RUCC type ranges from 1 to 9. The values were divided by 10 before using for regression analysis.)
18. Out trips: Average number of out-of-county trips of a county as provided by University of Maryland [6]. (Feature was normalized before usage)

## Feature selection methods

### Forward selection

The steps for the forward selection method are as follows:

1. Fix a significance level (0.05, in our case). Let *C* be set of the candidate features and *S* be the set of currently selected features.
2. Fit all possible regression models considering one feature at a time from *C* and the features from *S*.
3. Select the feature *c* ∈ *C* that has lowest *p*-value over the constructed models.
4. If the *p*-value of *c* is less than the fixed significance level, then add *c* to *S*, delete *c* from *C*. Repeat step 2 to 4. Otherwise finish the process and return *S*.

### Backward elimination

The steps for the forward selection method are as follows:

1. Fix a significance level (0.05, in our case). Let *C* be set of the candidate features.
2. Fit a model considering all features in *C*. Select the feature *c* ∈ *C* that has the highest *p*-value.
3. If the *p*-value of *c* is more than the fixed significance level, then delete *c* from *C*. Repeat steps 2 to 3. Otherwise finish the process and return *C*.

### Stepwise selection

The steps for stepwise selection method are as follows:

1. Fix a significance level (0.05, in our case). Let *C* be set of the candidate features and *S* be the set of currently selected features.
2. Fit all possible regression models considering one feature at a time from *C* and the features from *S*.
3. Select the feature *c* ∈ *C* that has lowest *p*-value over the constructed models.
4. If the *p*-value of *c* is less than the fixed significance level, then add *c* to *S*. Otherwise, finish the process and return *S*.
5. Fit a model considering all features in *S*. Select the feature *s* ∈ *S* that has the highest *p*-value.
6. If the *p*-value of *s* is more than the fixed significance level, then delete *s* from *S*. Repeat steps 5 to 6. Otherwise, go to next step
7. Set *C* = *C* − *S*
8. If *C* = ∅, finish the process and return *S*. Otherwise, go back to step 2 and repeat.

---

**Figure S3:**
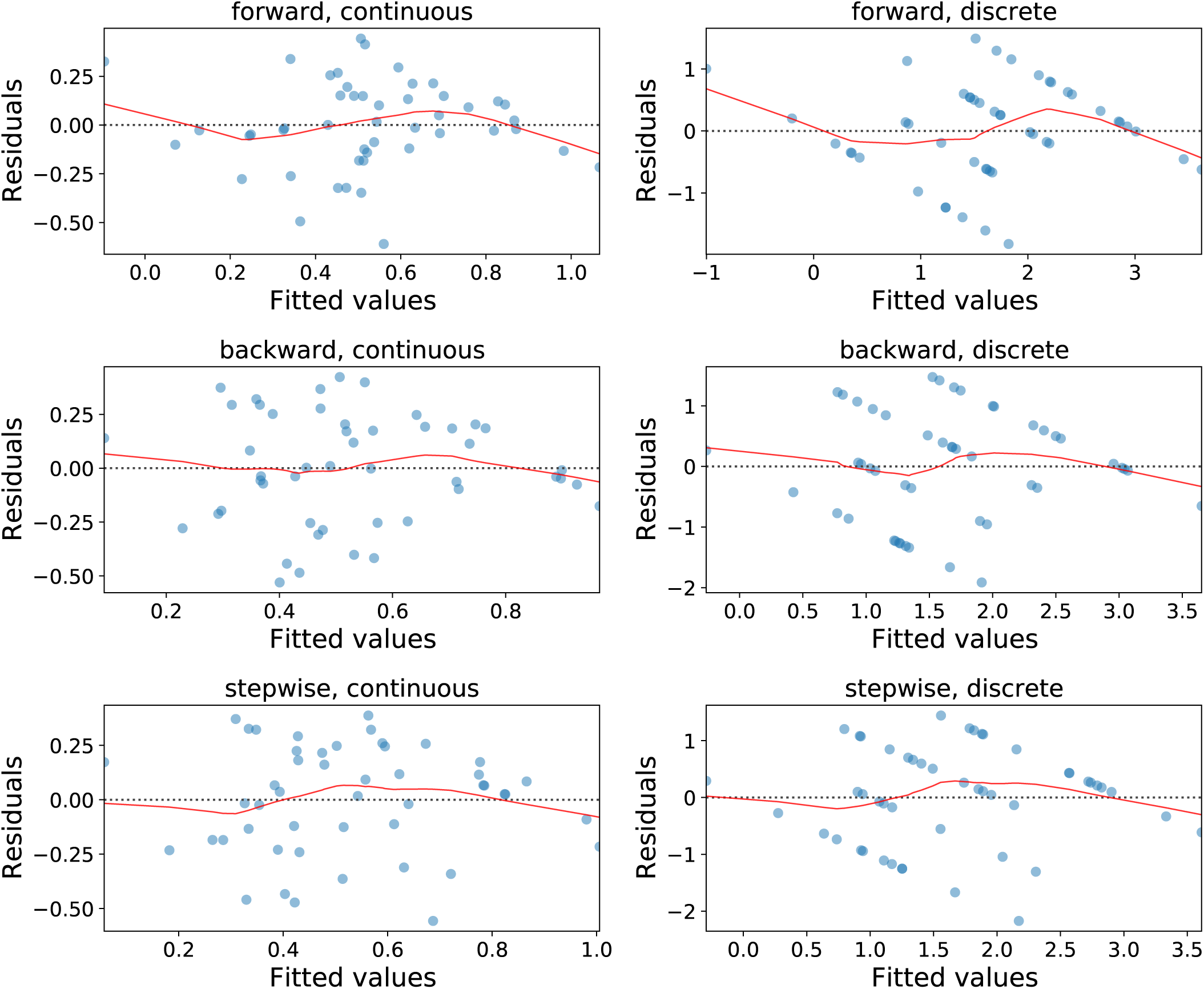
Residual vs fitted value plot for each linear regression model.

**Table S1:** Regression analyses results for all models

| Method | Target Variable Type |  |  |  |  |  |  |  |  |  |
| --- | --- | --- | --- | --- | --- | --- | --- | --- | --- | --- |
|  | Continuous |  |  |  |  | Discrete |  |  |  |  |
| Forward | **Multiple R<sup>2</sup>: 0.372**<br>**Adjusted R<sup>2</sup>: 0.329** |  |  |  |  | **Multiple R<sup>2</sup>: 0.423**<br>**Adjusted R<sup>2</sup>: 0.384** |  |  |  |  |
|  | **Variable** | **coef** | **std err** | **t** | **P>\|t\|** | **Variable** | **coef** | **std err** | **t** | **P>\|t\|** |
|  | Entropy of case rate | -0.4109 | 0.325 | -1.263 | 0.213 | Entropy of case rate | -0.7931 | 1.143 | -0.694 | 0.491 |
|  | University population | 1.1793 | 0.319 | 3.701 | 0.001 | University population | 4.9982 | 1.120 | 4.465 | 0.000 |
|  | Population | -0.0394 | 0.014 | -2.842 | 0.007 | Population | -0.1774 | 0.049 | -3.643 | 0.001 |
|  | const | 0.5810 | 0.293 | 1.981 | 0.054 | const | 1.1475 | 1.031 | 1.114 | 0.272 |
| Backward | **Multiple R<sup>2</sup>: 0.504**<br>**Adjusted R<sup>2</sup>: 0.445** |  |  |  |  | **Multiple R<sup>2</sup>: 0.585**<br>**Adjusted R<sup>2</sup>: 0.535** |  |  |  |  |
|  | **Variable** | **coef** | **std err** | **t** | **P>\|t\|** | **Variable** | **coef** | **std err** | **t** | **P>\|t\|** |
|  | Median Income | -0.946 | 0.334 | -2.832 | 0.007 | Median Income | -3.5303 | 1.121 | -3.150 | 0.003 |
|  | University population | 0.8010 | 0.311 | 2.574 | 0.014 | University population | 3.4611 | 1.044 | 3.315 | 0.002 |
|  | Entropy of Case rate | -0.6517 | 0.31 | -2.101 | 0.042 | Entropy of Case rate | -1.8552 | 1.041 | -1.702 | 0.082 |
|  | Population | -0.0342 | 0.015 | -2.339 | 0.024 | Population | -0.1679 | 0.049 | -3.420 | 0.001 |
|  | RUCC Type | -1.2749 | 0.449 | -2.841 | 0.007 | RUCC Type | -5.4985 | 1.505 | -3.652 | 0.001 |
|  | const | 1.7834 | 0.448 | 3.979 | 0.000 | const | 6.0309 | 1.504 | 4.010 | 0.000 |
| Stepwise | **Multiple R<sup>2</sup>: 0.412**<br>**Adjusted R<sup>2</sup>: 0.372** |  |  |  |  | **Multiple R<sup>2</sup>: 0.467**<br>**Adjusted R<sup>2</sup>: 0.430** |  |  |  |  |
|  | **Variable** | **coef** | **std err** | **t** | **P>\|t\|** | **Variable** | **coef** | **std err** | **t** | **P>\|t\|** |
|  | University population | 1.1746 | 0.295 | 3.975 | 0.000 | University population | 4.7873 | 1.032 | 4.641 | 0.000 |
|  | Population | -0.0452 | 0.012 | -3.833 | 0.000 | Population | -0.1852 | 0.041 | -4.495 | 0.000 |
|  | Entropy of interaction rate | -1.3402 | 0.618 | -2.169 | 0.213 | Entropy of interaction rate | -0.7931 | 1.143 | -0.694 | 0.491 |
|  | const | 1.4478 | 0.569 | 2.545 | 0.014 | const | 4.4395 | 1.986 | 2.236 | 0.031 |

**Figure S4:**
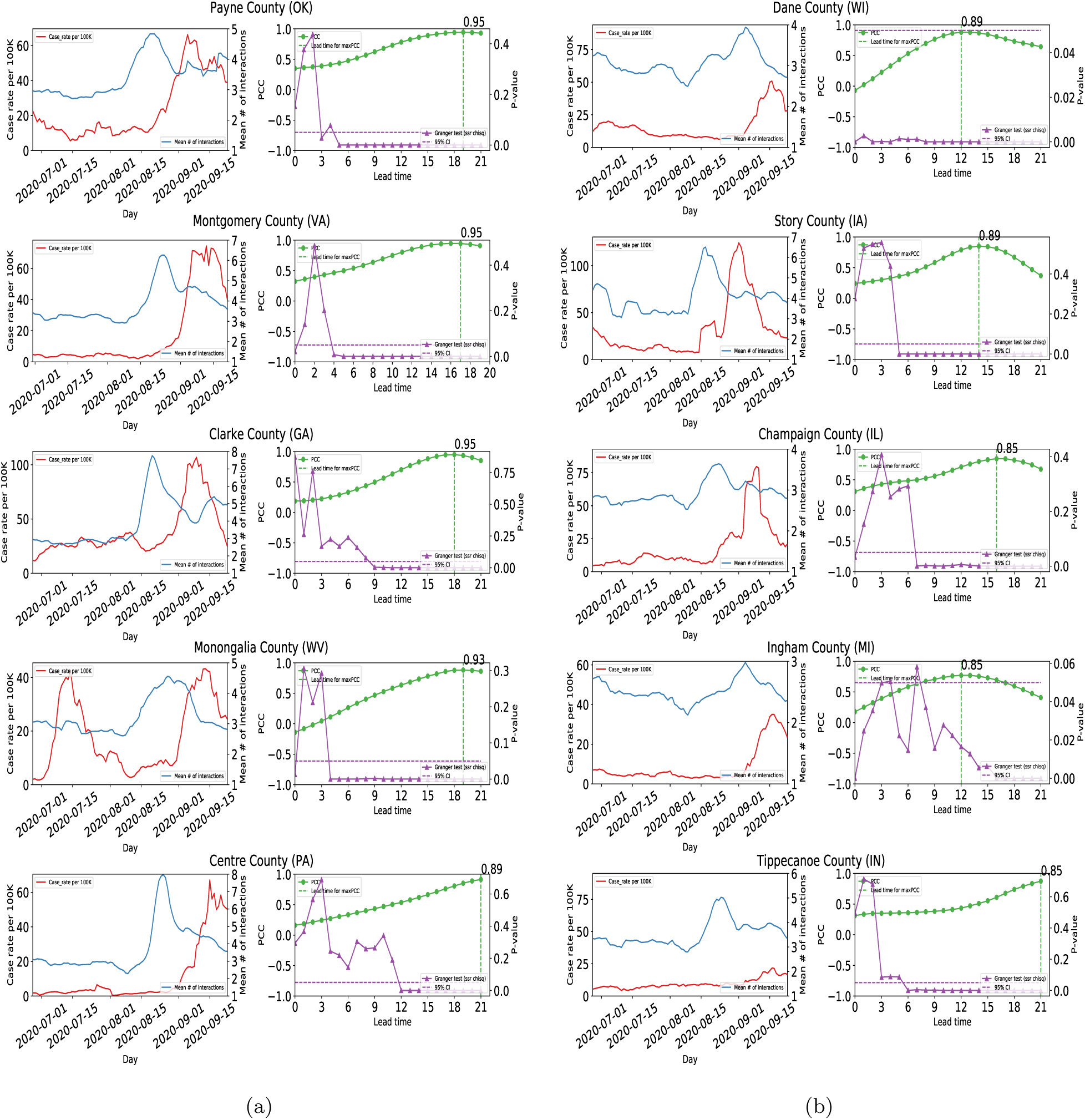
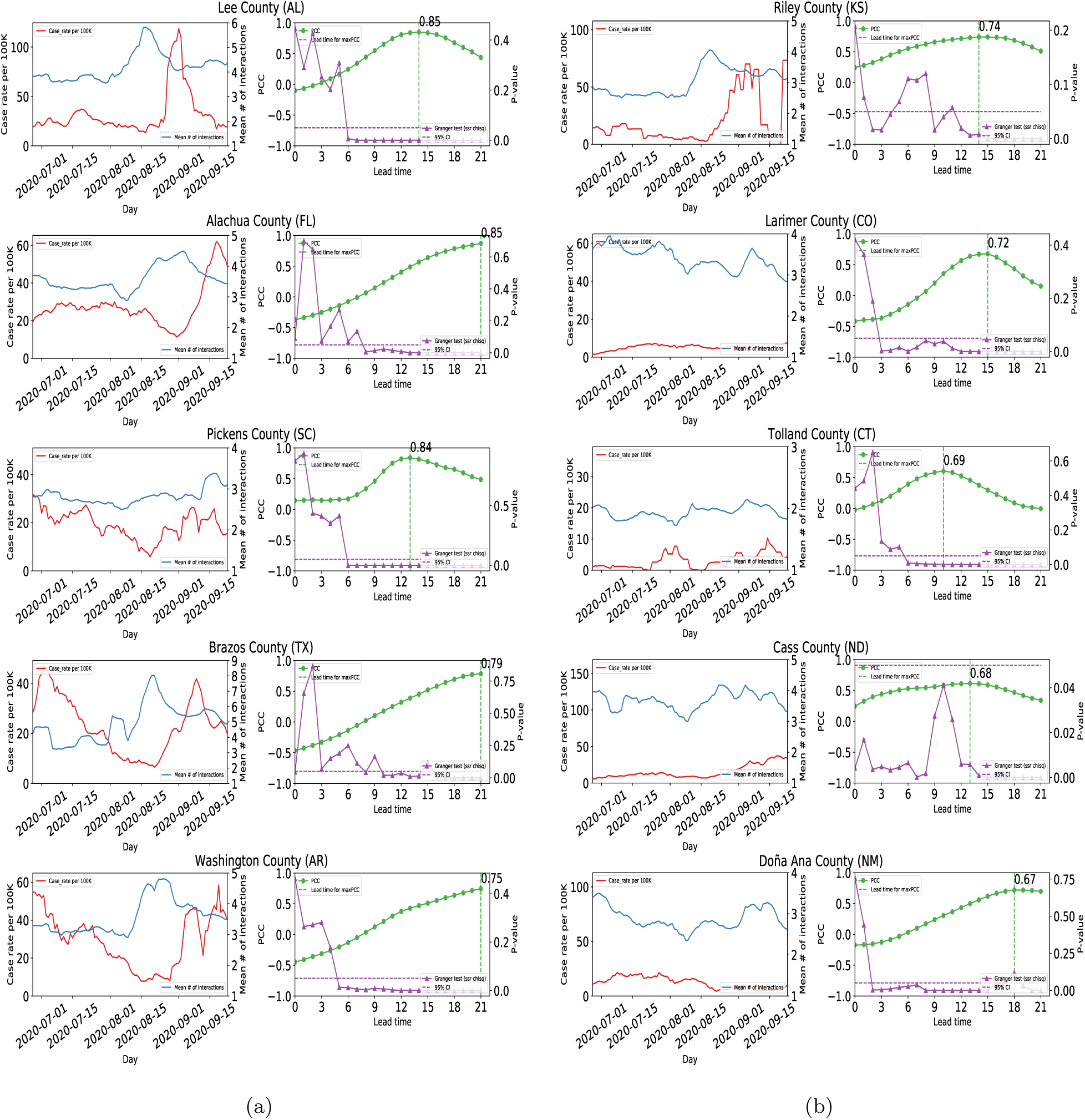
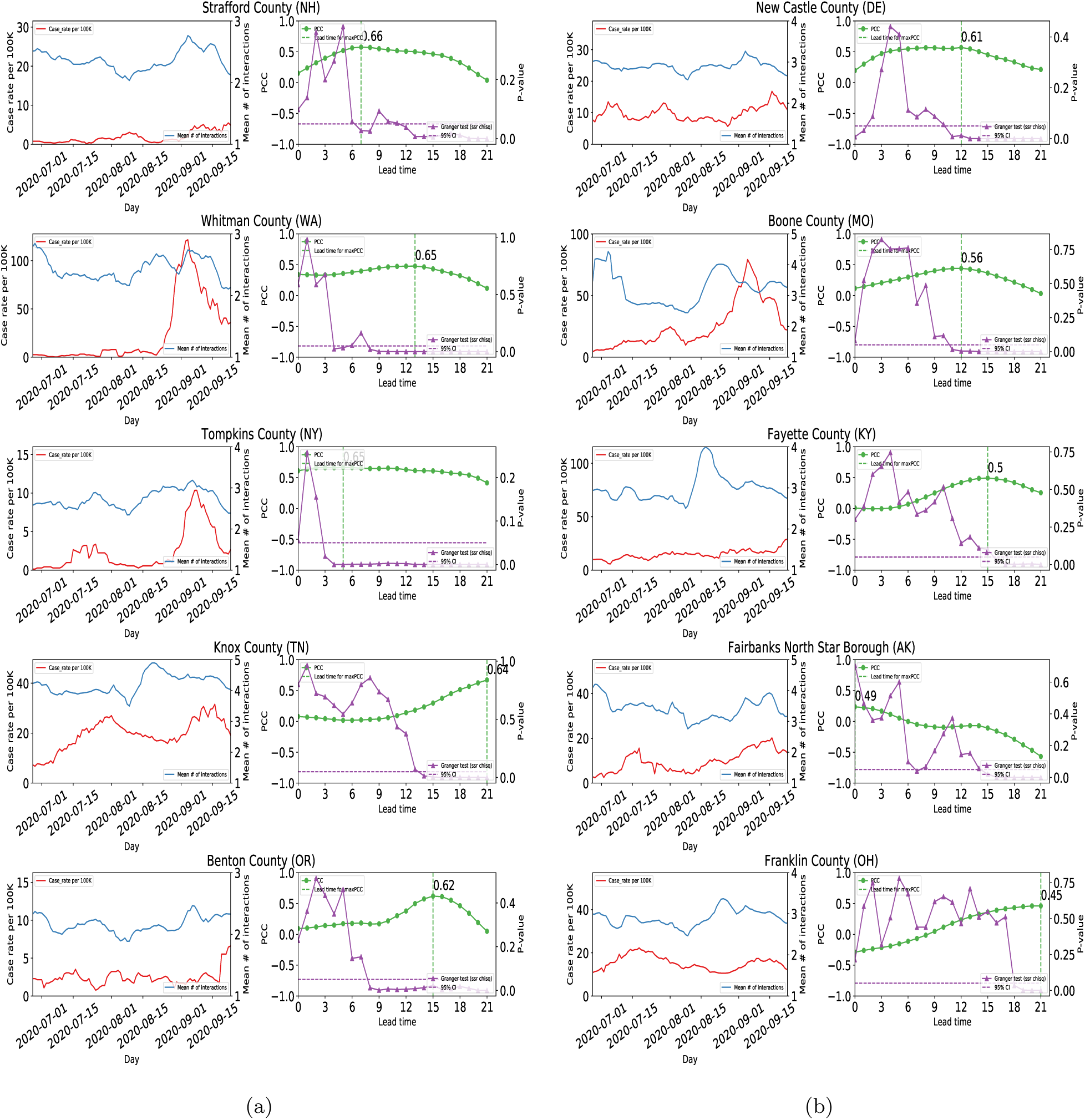
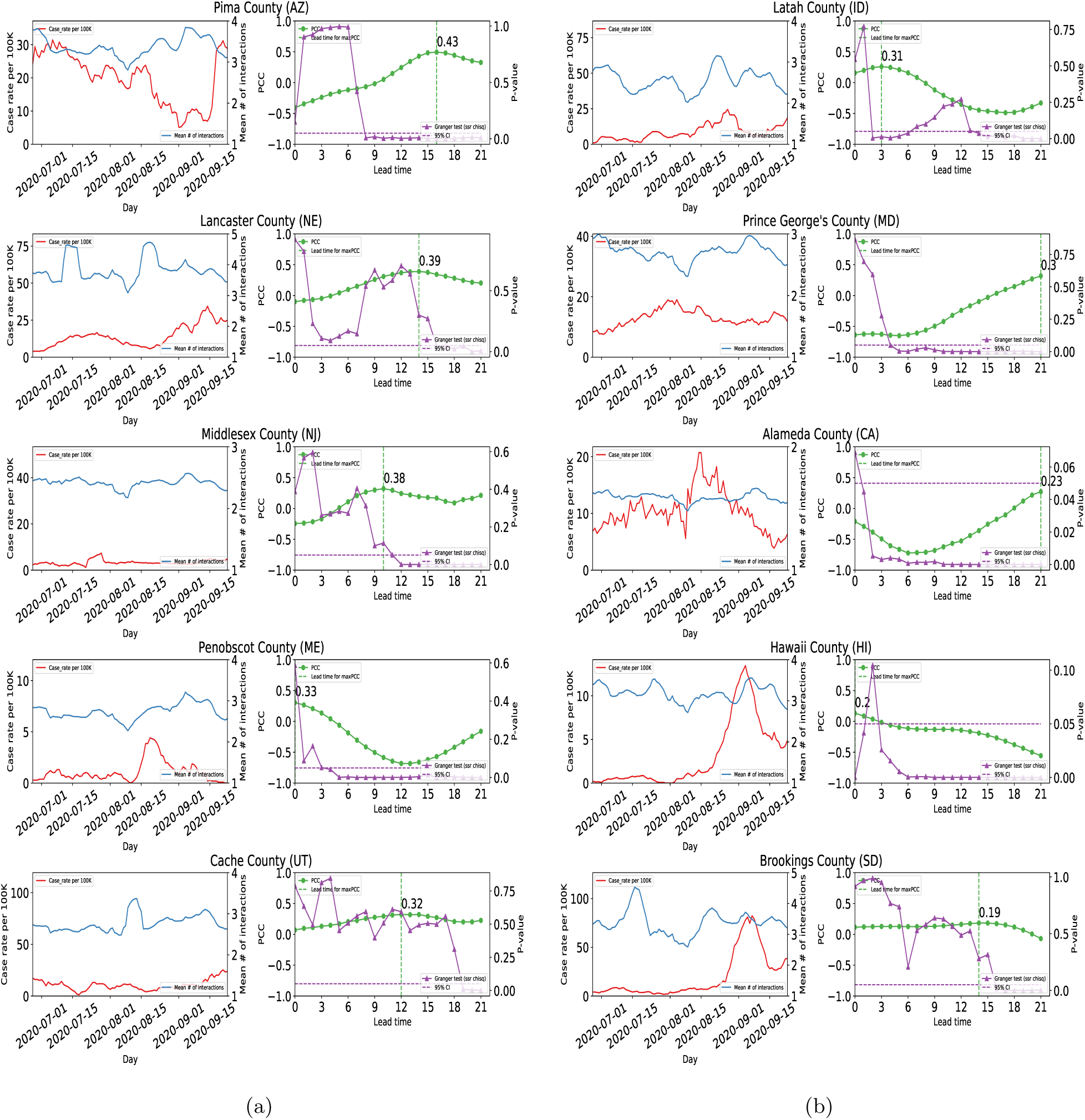
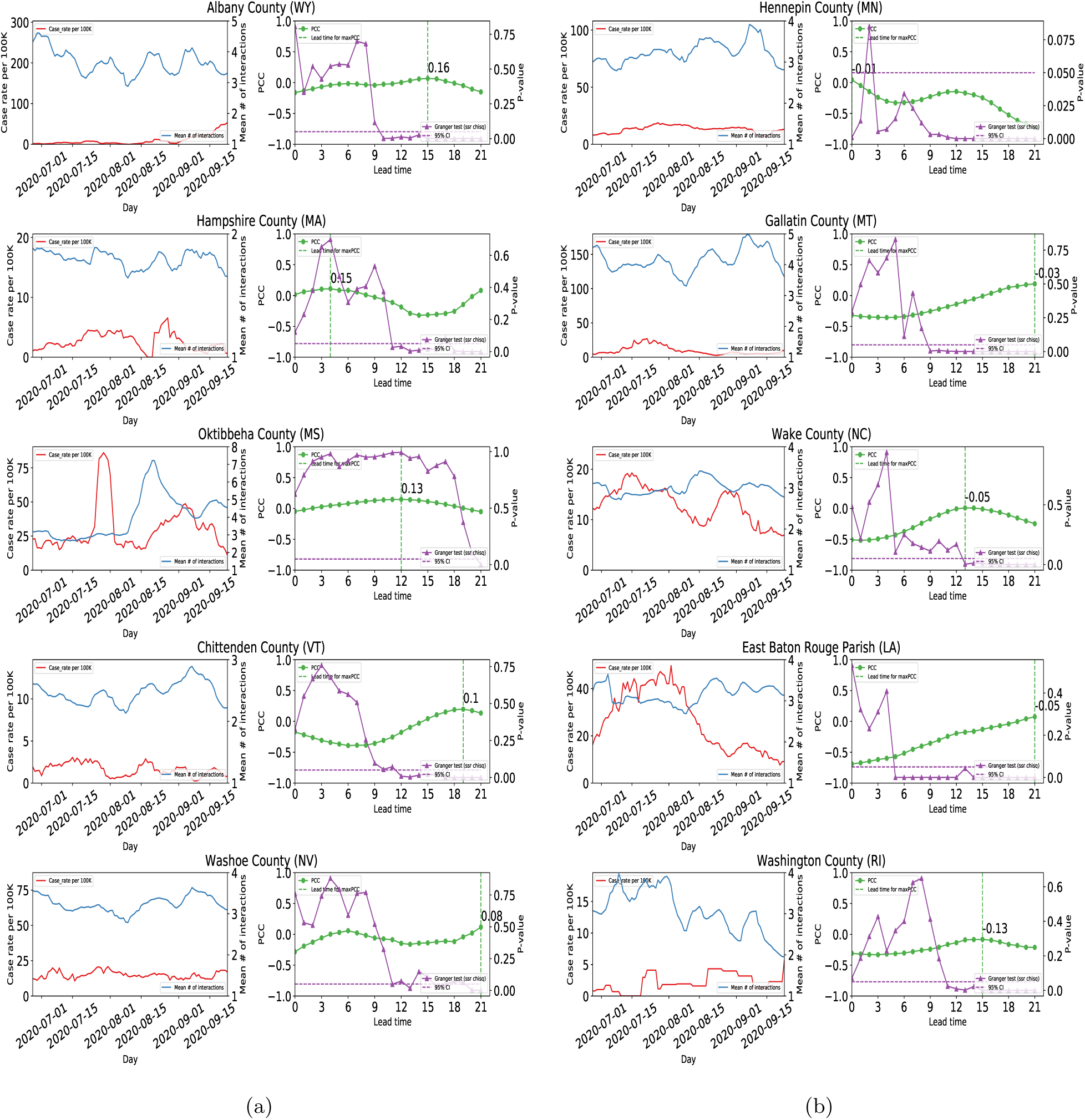
Results for all LGUCs.

**Table S2:** List of LGUCs and corresponding Universities with maxPCC and lead times.

| LGUC | University | maxPCC | Lead time |
| --- | --- | --- | --- |
| Payne, OK | Oklahoma State University | 0.95 | 19 |
| Montgomery, VA | Virginia Polytechnic University | 0.95 | 17 |
| Clarke, GA | University of Georgia | 0.95 | 18 |
| Monongalia, WV | West Virginia University | 0.93 | 18 |
| Centre, PA | Pennsylvania State University | 0.89 | 20 |
| Dane, WI | University of Wisconsin-Madison | 0.89 | 12 |
| Story, IA | Iowa State University | 0.89 | 14 |
| Alachua, FL | University of Florida | 0.85 | 20 |
| Ingham, MI | Michigan State University | 0.85 | 12 |
| Champaign, IL | University of Illinois at Urbana-Champaign | 0.85 | 16 |
| Lee, AL | Auburn University | 0.85 | 14 |
| Tippecanoe, IN | Purdue University | 0.85 | 20 |
| Pickens, SC | Clemson University | 0.84 | 13 |
| Brazos, TX | Texas A&M University | 0.79 | 20 |
| Washington, AR | University of Arkansas | 0.75 | 20 |
| Riley, KS | Kansas State University | 0.74 | 14 |
| Larimer, CO | Colorado State University | 0.72 | 14 |
| Tolland, CT | University of Connecticut | 0.69 | 10 |
| Cass, ND | North Dakota State University | 0.68 | 13 |
| Doña Ana, NM | New Mexico State University | 0.67 | 20 |
| Strafford, NH | University of New Hampshire | 0.66 | 7 |
| Whitman, WA | Washington State University | 0.65 | 13 |
| Tompkins, NY | Cornell University | 0.65 | 5 |
| Knox, TN | University of Tennessee | 0.64 | 20 |
| Benton, OR | Oregon State University | 0.62 | 15 |
| New Castle, DE | University of Delaware | 0.61 | 12 |
| Boone, MO | University of Missouri | 0.56 | 11 |
| Fayette, KY | University of Kentucky | 0.50 | 15 |
| Fairbanks North Star, AK | University of Alaska Fairbanks | 0.49 | 0 |
| Franklin, OH | Ohio State University | 0.45 | 20 |
| Pima, AZ | University of Arizona | 0.43 | 16 |
| Lancaster, NE | University of Nebraska-Lincoln | 0.39 | 14 |
| Middlesex, NJ | Rutgers University | 0.38 | 10 |
| Penobscot, ME | University of Maine | 0.33 | 0 |
| Cache, UT | Utah State University | 0.32 | 12 |
| Latah, ID | University of Idaho | 0.31 | 3 |
| Prince George's, MD | University of Maryland, College Park | 0.30 | 20 |
| Alameda, CA | University of California Berkeley | 0.23 | 20 |
| Hawaii, HI | University of Hawaii | 0.20 | 0 |
| Brookings, SD | South Dakota State University | 0.19 | 14 |
| Albany, WY | University of Wyoming | 0.16 | 15 |
| Hampshire, MA | University of Massachusetts Amherst | 0.15 | 3 |
| Oktibbeha, MS | Mississippi State University | 0.13 | 12 |
| Chittenden, VT | University of Vermont | 0.10 | 18 |
| Washoe, NV | University of Nevada, Reno | 0.08 | 20 |
| Hennepin, MN | University of Minnesota | -0.01 | 0 |
| Gallatin, MT | Montana State University | -0.03 | 18 |
| Wake, NC | North Carolina State University | -0.05 | 13 |
| East Baton Rouge, LA | Louisiana State University | -0.05 | 20 |
| Washington, RI | University of Rhode Island | -0.13 | 15 |

**Table S3:**
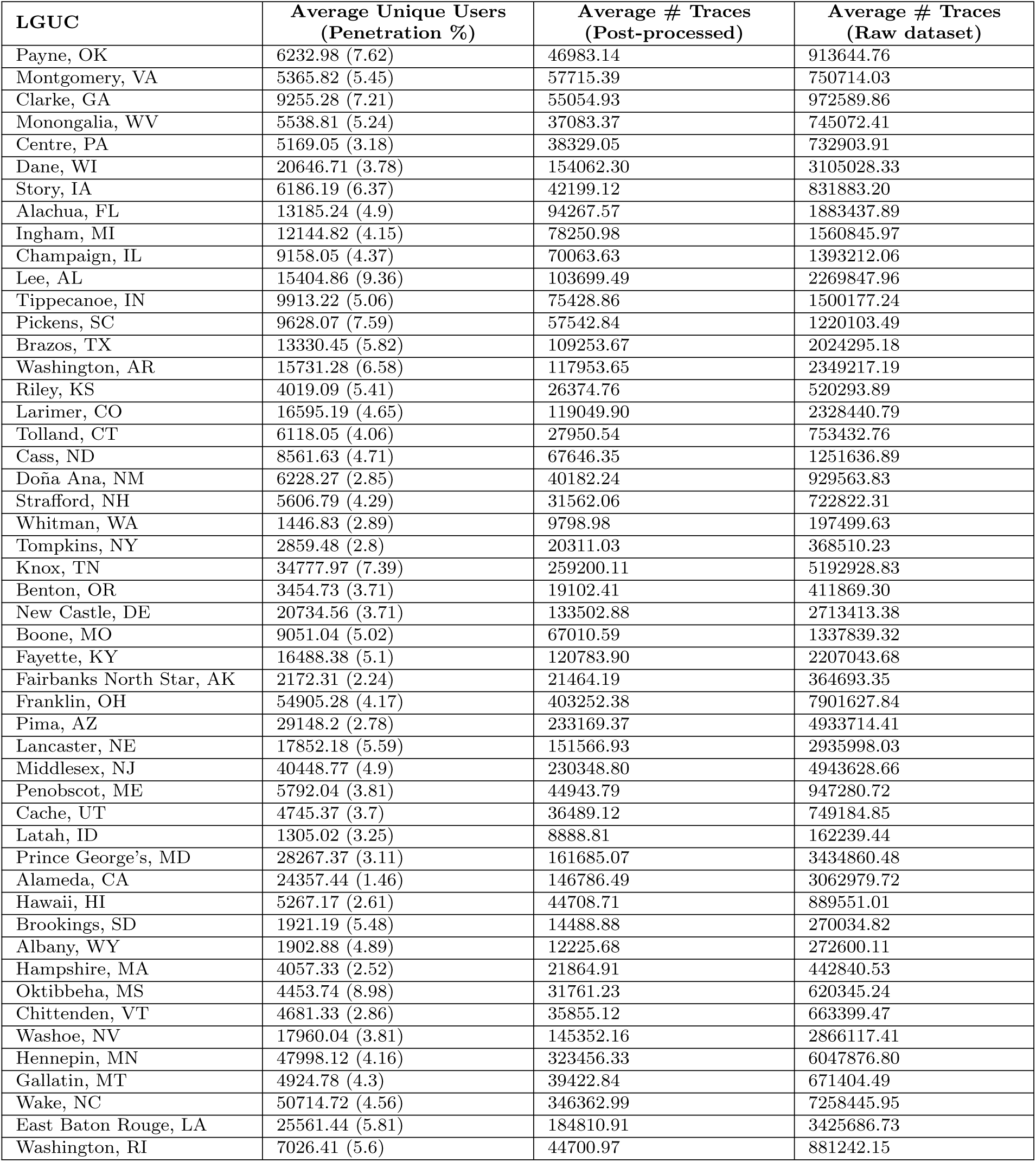
Demographics and app coverage statistics for X-Mode data. The second column is the average number of unique users in our Dataset for each day along with the penetration percentage, which is the percentage ratio of unique users to the population of the LGUC. The third and fourth column represent the number of traces in our dataset after and before stop point detection, respectively.

| LGUC | Average Unique Users<br>(Penetration %) | Average # Traces<br>(Post-processed) | Average # Traces<br>(Raw dataset) |
| --- | --- | --- | --- |
| Payne, OK | 6232.98 (7.62) | 46983.14 | 913644.76 |
| Montgomery, VA | 5365.82 (5.45) | 57715.39 | 750714.03 |
| Clarke, GA | 9255.28 (7.21) | 55054.93 | 972589.86 |
| Monongalia, WV | 5538.81 (5.24) | 37083.37 | 745072.41 |
| Centre, PA | 5169.05 (3.18) | 38329.05 | 732903.91 |
| Dane, WI | 20646.71 (3.78) | 154062.30 | 3105028.33 |
| Story, IA | 6186.19 (6.37) | 42199.12 | 831883.20 |
| Alachua, FL | 13185.24 (4.9) | 94267.57 | 1883437.89 |
| Ingham, MI | 12144.82 (4.15) | 78250.98 | 1560845.97 |
| Champaign, IL | 9158.05 (4.37) | 70063.63 | 1393212.06 |
| Lee, AL | 15404.86 (9.36) | 103699.49 | 2269847.96 |
| Tippecanoe, IN | 9913.22 (5.06) | 75428.86 | 1500177.24 |
| Pickens, SC | 9628.07 (7.59) | 57542.84 | 1220103.49 |
| Brazos, TX | 13330.45 (5.82) | 109253.67 | 2024295.18 |
| Washington, AR | 15731.28 (6.58) | 117953.65 | 2349217.19 |
| Riley, KS | 4019.09 (5.41) | 26374.76 | 520293.89 |
| Larimer, CO | 16595.19 (4.65) | 119049.90 | 2328440.79 |
| Tolland, CT | 6118.05 (4.06) | 27950.54 | 753432.76 |
| Cass, ND | 8561.63 (4.71) | 67646.35 | 1251636.89 |
| Doña Ana, NM | 6228.27 (2.85) | 40182.24 | 929563.83 |
| Strafford, NH | 5606.79 (4.29) | 31562.06 | 722822.31 |
| Whitman, WA | 1446.83 (2.89) | 9798.98 | 197499.63 |
| Tompkins, NY | 2859.48 (2.8) | 20311.03 | 368510.23 |
| Knox, TN | 34777.97 (7.39) | 259200.11 | 5192928.83 |
| Benton, OR | 3454.73 (3.71) | 19102.41 | 411869.30 |
| New Castle, DE | 20734.56 (3.71) | 133502.88 | 2713413.38 |
| Boone, MO | 9051.04 (5.02) | 67010.59 | 1337839.32 |
| Fayette, KY | 16488.38 (5.1) | 120783.90 | 2207043.68 |
| Fairbanks North Star, AK | 2172.31 (2.24) | 21464.19 | 364693.35 |
| Franklin, OH | 54905.28 (4.17) | 403252.38 | 7901627.84 |
| Pima, AZ | 29148.2 (2.78) | 233169.37 | 4933714.41 |
| Lancaster, NE | 17852.18 (5.59) | 151566.93 | 2935998.03 |
| Middlesex, NJ | 40448.77 (4.9) | 230348.80 | 4943628.66 |
| Penobscot, ME | 5792.04 (3.81) | 44943.79 | 947280.72 |
| Cache, UT | 4745.37 (3.7) | 36489.12 | 749184.85 |
| Latah, ID | 1305.02 (3.25) | 8888.81 | 162239.44 |
| Prince George's, MD | 28267.37 (3.11) | 161685.07 | 3434860.48 |
| Alameda, CA | 24357.44 (1.46) | 146786.49 | 3062979.72 |
| Hawaii, HI | 5267.17 (2.61) | 44708.71 | 889551.01 |
| Brookings, SD | 1921.19 (5.48) | 14488.88 | 270034.82 |
| Albany, WY | 1902.88 (4.89) | 12225.68 | 272600.11 |
| Hampshire, MA | 4057.33 (2.52) | 21864.91 | 442840.53 |
| Oktibbeha, MS | 4453.74 (8.98) | 31761.23 | 620345.24 |
| Chittenden, VT | 4681.33 (2.86) | 35855.12 | 663399.47 |
| Washoe, NV | 17960.04 (3.81) | 145352.16 | 2866117.41 |
| Hennepin, MN | 47998.12 (4.16) | 323456.33 | 6047876.80 |
| Gallatin, MT | 4924.78 (4.3) | 39422.84 | 671404.49 |
| Wake, NC | 50714.72 (4.56) | 346362.99 | 7258445.95 |
| East Baton Rouge, LA | 25561.44 (5.81) | 184810.91 | 3425686.73 |
| Washington, RI | 7026.41 (5.6) | 44700.97 | 881242.15 |

**Table S4:** List of land grant universities with corresponding US counties.

| LGUC | University | LGUC<br>Population<br>(in 100K) | University<br>Population<br>(in 100K) |
| --- | --- | --- | --- |
| Payne, OK | Oklahoma State University | 0.81784 | 0.27200 |
| Montgomery, VA | Virginia Polytechnic University | 0.98535 | 0.36882 |
| Clarke, GA | University of Georgia | 1.28331 | 0.42708 |
| Monongalia, WV | West Virginia University | 1.05612 | 0.29323 |
| Centre, PA | Pennsylvania State University | 1.62385 | 0.49935 |
| Dane, WI | University of Wisconsin-Madison | 5.46695 | 0.47392 |
| Story, IA | Iowa State University | 0.97117 | 0.37944 |
| Alachua, FL | University of Florida | 2.69043 | 0.58749 |
| Ingham, MI | Michigan State University | 2.92406 | 0.55332 |
| Champaign, IL | University of Illinois at Urbana-Champaign | 2.09689 | 0.54985 |
| Lee, AL | Auburn University | 1.64542 | 0.32442 |
| Tippecanoe, IN | Purdue University | 1.95732 | 0.47412 |
| Pickens, SC | Clemson University | 1.26884 | 0.28310 |
| Brazos, TX | Texas A&M University | 2.29211 | 0.73267 |
| Washington, AR | University of Arkansas | 2.39187 | 0.30459 |
| Riley, KS | Kansas State University | 0.74232 | 0.24546 |
| Larimer, CO | Colorado State University | 3.56899 | 0.37356 |
| Tolland, CT | University of Connecticut | 1.50721 | 0.29683 |
| Cass, ND | North Dakota State University | 1.81923 | 0.14503 |
| Doña Ana, NM | New Mexico State University | 2.18195 | 0.16143 |
| Strafford, NH | University of New Hampshire | 1.30633 | 0.16673 |
| Whitman, WA | Washington State University | 0.50104 | 0.35149 |
| Tompkins, NY | Cornell University | 1.02180 | 0.24189 |
| Knox, TN | University of Tennessee | 4.70313 | 0.31477 |
| Benton, OR | Oregon State University | 0.93053 | 0.38110 |
| New Castle, DE | University of Delaware | 5.58753 | 0.25885 |
| Boone, MO | University of Missouri | 1.80463 | 0.33333 |
| Fayette, KY | University of Kentucky | 3.23152 | 0.30981 |
| Fairbanks North Star, AK | University of Alaska Fairbanks | 0.96849 | 0.12297 |
| Franklin, OH | Ohio State University | 13.16756 | 0.66178 |
| Pima, AZ | University of Arizona | 10.47279 | 0.48653 |
| Lancaster, NE | University of Nebraska-Lincoln | 3.19090 | 0.28686 |
| Middlesex, NJ | Rutgers University | 8.25062 | 0.57162 |
| Penobscot, ME | University of Maine | 1.52148 | 0.13091 |
| Cache, UT | Utah State University | 1.28289 | 0.32963 |
| Latah, ID | University of Idaho | 0.40108 | 0.13956 |
| Prince George's, MD | University of Maryland, College Park | 9.09327 | 0.44486 |
| Alameda, CA | University of California Berkeley | 16.71329 | 0.45183 |
| Hawaii, HI | University of Hawaii | 2.01513 | 0.20653 |
| Brookings, SD | South Dakota State University | 0.35077 | 0.14785 |
| Albany, WY | University of Wyoming | 0.38880 | 0.13926 |
| Hampshire, MA | University of Massachusetts Amherst | 1.60830 | 0.35116 |
| Oktibbeha, MS | Mississippi State University | 0.49587 | 0.24251 |
| Chittenden, VT | University of Vermont | 1.63774 | 0.15698 |
| Washoe, NV | University of Nevada, Reno | 4.71519 | 0.24246 |
| Hennepin, MN | University of Minnesota-Twin Cities | 11.5242 | 0.61787 |
| Gallatin, MT | Montana State University | 1.14434 | 0.18898 |
| Wake, NC | North Carolina State University | 11.11761 | 0.39822 |
| East Baton Rouge, LA | Louisiana State University | 4.40059 | 0.33758 |
| Washington, RI | University of Rhode Island | 1.25577 | 0.20628 |

## References

[1] Economic Research Service, United States Department of Agriculture. https://www.ers.usda.gov/data-products/county-level-data-sets/download-data/, visited 2020-11-10.

[2] U.S. coronavirus cases and deaths. https://usafacts.org/visualizations/coronavirus-covid-19-spread-map/, visited 2020-11-10.

[3] X-Mode. https://xmode.io/, visited 2020-11-10.

[4] Samuel Altmann, Luke Milsom, Hannah Zillessen, Raffaele Blasone, Frederic Gerdon, Ruben Bach, Frauke Kreuter, Daniele Nosenzo, Severine Toussaert, and Johannes Abeler. Acceptability of app-based contact tracing for COVID-19: Cross-country survey evidence. Available at SSRN 3590505, 2020.

[5] Martin S. Andersen, Ana I. Bento, Anirban Basu, Chris Marsicano, and Kosali Simon. College openings, mobility, and the incidence of covid-19 cases. medRxiv, 2020.

[6] Hamada S. Badr, Hongru Du, Maximilian Marshall, Ensheng Dong, Marietta M. Squire, and Lauren M. Gardner. Association between mobility patterns and COVID-19 transmission in the USA: a mathematical modelling study. The Lancet Infectious Diseases, 20(11):1247–1254, 2020.

[7] U.S. Census Bureau. TIGER/Line shapefiles. https://www.census.gov/geographies/mapping-files/time-series/geo/tiger-line-file.html, visited 2020-11-10.

[8] Davidson College. College Crisis Initiative. https://collegecrisis.shinyapps.io/dashboard.

[9] Benjamin D. Dalziel, Stephen Kissler, Julia R. Gog, Cecile Viboud, Ottar N. Bjørnstad, C. Jessica E. Metcalf, and Bryan T. Grenfell. Urbanization and humidity shape the intensity of influenza epidemics in US cities. Science, 362(6410):75–79, 2018.

[10] Economic Research Service, United States Department of Agriculture. Rural Urban Continuum Codes. https://www.ers.usda.gov/data-products/rural-urban-continuum-codes.aspx, visited 2020-11-10.

[11] Nuria Oliver, Bruno Lepri, Harald Sterly, Renaud Lambiotte, Sébastien Deletaille, Marco De Nadai, Emmanuel Letouzé, Albert Ali Salah, Richard Benjamins, Ciro Cattuto, Vittoria Colizza, Nicolas de Cordes, Samuel P. Fraiberger, Till Koebe, Sune Lehmann, Juan Murillo, Alex Pentland, Phuong N Pham, Frédéric Pivetta, Jari Saramäki, Samuel V. Scarpino, Michele Tizzoni, Stefaan Verhulst, and Patrick Vinck. Mobile phone data for informing public health actions across the COVID-19 pandemic life cycle. Science Advances, 6(23), 2020.

[12] Luca Pappalardo, F. Simini, G. Barlacchi, and R. Pellungrini. scikit-mobility: A Python library for the analysis, generation and risk assessment of mobility data. 1907.07062, 2019.

[13] Lior Rennert, Corey Andrew Kalbaugh, Lu Shi, and Christopher McMahan. Reopening universities during the COVID-19 pandemic: A testing strategy to minimize active cases and delay outbreaks. medRxiv:2020.07.06.20147272, 2020.

[14] Pradeep Sahu. Closure of universities due to Coronavirus Disease 2019 (COVID-19): Impact on education and mental health of students and academic staff. Cureus, 12(4), 2020.

[15] Skipper Seabold and Josef Perktold. statsmodels: Econometric and statistical modeling with Python. In 9th Python in Science Conference, 2010.

[16] The New York Times. Tracking the Coronavirus at U.S. Colleges and Universities. https://www.nytimes.com/interactive/2020/us/covid-college-cases-tracker.html.

## References

[2] U.S. Census Bureau. Population and housing unit estimates. https://www.census.gov/programs-surveys/popest/technical-documentation/file-layouts.html, visited 2020-11-10.

[3] Economic Research Service, United States Department of Agriculture. Rural Urban Continuum Codes. https://www.ers.usda.gov/data-products/rural-urban-continuum-codes.aspx, visited 2020-11-10.

[4] National Center for Education Statistics. IPEDS. https://nces.ed.gov/ipeds/use-the-data, visited 2020-11-10.

[5] Luca Pappalardo, F. Simini, G. Barlacchi, and R. Pellungrini. scikit-mobility: A Python library for the analysis, generation and risk assessment of mobility data. 1907.07062, 2019.

[6] Lei Zhang, Sepehr Ghader, Michael L Pack, Chenfeng Xiong, Aref Darzi, Mofeng Yang, Qianqian Sun, AliAkbar Kabiri, and Songhua Hu. An interactive covid-19 mobility impact and social distancing analysis platform. medRxiv:2020.04.29.20085472, 2020.

